# Neutrophil Extracellular Trap Biomarker Titers and Delayed Cerebral Ischemia in Patients With Aneurysmal Subarachnoid Hemorrhage

**DOI:** 10.1101/2022.02.01.22270255

**Authors:** Jens Witsch, Valérie Spalart, Kimberly Martinod, Hauke Schneider, Joachim Oertel, Jürgen Geisel, Philipp Hendrix, Sina Hemmer

**Affiliations:** Department of Neurology, University of Pennsylvania School of Medicine, Philadelphia, Pennsylvania, USA; Center for Molecular and Vascular Biology, Department of Cardiovascular Sciences, KU Leuven, Leuven, Belgium; Department of Neurology, University Hospital Augsburg, Augsburg, Germany; Department of Neurosurgery, Saarland University Medical Center, Homburg/Saar, Germany; Department of Clinical Chemistry and Laboratory Medicine, Saarland University Medical Center, Homburg/Saar, Germany

## Abstract

**Background:** Myeloperoxidase (MPO)-DNA complexes, biomarkers of neutrophil extracellular traps, have been associated with arterial and venous thrombosis. Their role in patients with aneurysmal subarachnoid hemorrhage (aSAH) is unknown. Here, we sought to explore whether serum MPO-DNA-complexes are present in patients with aSAH, and whether these levels are associated with delayed cerebral ischemia (DCI).

**Methods:** We performed a post-hoc analysis of a prospective, blinded, observational single-center biomarker study that enrolled consecutive patients with spontaneous SAH between July 2018 and September 2020. Serum samples obtained on admission and on hospital day 4 were analyzed for concentrations of MPO-DNA complexes. The primary outcome was the occurrence of DCI defined as new infarction on brain CT-scan. The secondary outcome was clinical vasospasm, a composite of clinical and transcranial doppler parameters. We used Wilcoxon’s signed-rank-test to assess for differences between paired measures.

**Results:** Among 100 patients with spontaneous SAH, mean age 59 (SD±13) years, 55% women, 78 patients had an aneurysmal SAH and complete DCI information. Among these, 29 (37%) developed DCI. MPO-DNA complexes were detected in all samples. The median MPO-DNA level was 33 ng/ml (IQR, 18-43 ng/ml) on admission, and 22 ng/ml (IQR, 11-31 ng/ml) on day 4 (Mann-Whitney test: p=0.015). In the primary outcome analysis, we found a significant reduction in MPO-DNA levels from admission to day 4 in patients with DCI (paired test, p=0.036) but not in those without DCI (p=0.171). The secondary analysis showed a similar reduction in MPO-DNA levels between admission and day 4 in patients with (p=0.006), but not in those without clinical vasospasm (p=0.473).

**Conclusions:** MPO-DNA-complexes are detectable in peripheral serum samples of patients with aSAH. A pronounced reduction in MPO-DNA levels over the first days after aSAH might herald DCI. The potential of MPO-DNA levels to serve as a biomarker of DCI requires further exploration.

---

Aneurysmal subarachnoid hemorrhage (SAH) is a severe form of hemorrhagic stroke, associated with 18% in-hospital mortality, and about 30% poor long-term outcome.^1,2^ In about one third of cases the hospitalization is complicated by delayed cerebral ischemia (DCI), the occurrence of secondary ischemic infarcts, identified on brain imaging.^3^ Most DCI occurs 4 to 14 days after the initial hemorrhage and is strongly associated with poor outcome.^4,5^ However, effective prediction and prevention measures of DCI are lacking.^5^ While DCI development might be preceded by clinical deterioration, angiographic vasospasm, increase in transcranial doppler flow velocities, or electroencephalographic changes, DCI often occurs silently and is mostly identified in retrospect.^6-9^ Finding markers of impending DCI could lead to a paradigm shift in the diagnosis and possibly in the prevention and treatment of this severe complication of SAH.^10^ Neutrophil extracellular traps (NETs) have been described as a central mechanism of arterial and venous thrombus formation.^11^ Since their definition in 2004 as a neutrophil innate immune mechanism,^12^ they have also been detrimentally associated with ischemic stroke, deep vein thrombosis, as well as immunothrombosis driven by coronavirus disease 2019 (COVID-19).^13-15^ NETs are the product of neutrophil hyperactivation leading to the release of extracellular “net-like” structures consisting of chromatin lined with neutrophil proteins. When released into the blood, they facilitate thrombus formation by promoting coagulation and binding platelets and other blood components. NETs have been visualized using electron microscopy^12^, and components of NETs can be detected in plasma of patients with acute thrombotic processes.^12^ Moreover, NETs can be therapeutically targeted by degradation with DNases, as demonstrated in animal studies^16-18^ and ex vivo thrombolysis experiments.^13,19^ While excess inflammation and increased neutrophil lymphocyte ratios have been linked to DCI and outcome after aSAH, only limited data is available on the detection of NETs in patients with SAH.^20-22^

In this exploratory study we focused on one major NET biomarker, myeloperoxidase (MPO)-DNA complexes. We leveraged prospective data from a cohort of patients with SAH undergoing prospective neurological and transcranial Doppler (TCD) exams, as well as standardized DCI assessment. We aimed to test two hypotheses: (1) MPO-DNA complexes can be detected in peripheral serum of patients with aSAH. (2) Changes in MPO-DNA-complex levels between admission and hospital day 4 are associated with the occurrence of DCI.

## Methods

### Study design and population

We conducted a post-hoc analysis of the high mobility group box 1 in aneurysmal subarachnoid hemorrhage (HIMOBASH) study, a prospective, blinded, single tertiary care center biomarker observational study designed to explore the role of high mobility group box 1 protein as a biomarker in aneurysmal subarachnoid hemorrhage.^23^ All clinical data for the present study were collected at Saarland University Medical Center, Germany; all laboratory analyses were conducted at KU Leuven, Belgium. For the HIMOBASH study, consecutive patients with spontaneous subarachnoid hemorrhage, 18 years of age or older, admitted to the neurosurgical intensive care unit at Saarland University Medical Center, Germany, were evaluated for study eligibility. The diagnosis of SAH was established through an admission brain CT. An aneurysmal etiology of SAH was confirmed through CT angiography or digital subtraction angiography (DSA). If DSA was negative for an aneurysmal bleeding source, patients underwent additional brain MRI and repeat-DSA after 7–10 days to rule out other pathologies and establish the diagnosis of non-aneurysmal SAH.

### Clinical management

Patients were treated according to current guidelines including monitoring in a dedicated neurointensive care unit, aneurysm treatment via clipping or coiling no later than 24 hours from admission, treatment of hydrocephalus via insertion of an external ventricular drain, and oral nimodipine administration. Bedside TCD studies were performed twice daily and additionally as required to corroborate suspected vasospasm.^23^ If a delayed neurological deficit (DND) was identified (as defined in the *Outcomes* paragraph), hyperdynamic therapy was initiated avoiding hypovolemia and escalating the mean arterial blood pressure levels using vasopressors. If vasospasm was suspected, decisions on additional imaging or neurointerventional rescue therapies were made on a case-by-case basis after reaching interdisciplinary consensus.

### Outcomes

#### Delayed cerebral ischemia (DCI)

DCI was defined as a new ischemic infarct on brain imaging that was not present on the routine post-clipping/-coiling CT brain one day after aneurysm treatment. Patients who died before another follow-up CT scan was obtained, were deemed ineligible for DCI assessment, and were thus excluded from this study.

#### Clinical vasospasm

Clinical vasospasm was defined as a composite outcome consisting of at least one of the two following occurrences: 1) the diagnosis of DND; 2) abnormal TCD findings. ***DND***. The definition of DND followed the consensus statement of the Neurocritical Care Society.^24^ Glasgow Coma Scale (GCS) scores were assessed hourly for each patient. DND was diagnosed if a reduction of ≥2 on the GCS occurred or if a new focal neurological deficit was identified while other potential causes were eliminated.

#### Transcranial doppler studies

Bedside TCD were conducted at least twice daily to diagnose suspected vasospasm. The following TCD criteria were considered indicative of potential vasospasm: a middle cerebral artery (MCA) peak systolic velocity (PSV) of ≥ 120 cm/s that was not attributable to generalized hyperperfusion, a peak systolic velocity increase of ≥ 50 cm/s compared to the admission TCD exam, or a Lindegaard ratio (mean flow velocity in the MCA divided by mean flow velocity in the ipsilateral extracranial internal carotid artery) of ≥ 3. Isolated angiographic vasospasm – although assessed in the original HIMOBASH cohort – was a priori excluded as an endpoint for the purpose of this study for its poor correlation with DCI.^23^

##### Serum collection and Laboratory Measurements

For laboratory analysis of MPO-DNA-complex concentrations, we used serum samples collected on admission (day 0) and on day 4, as previously described.^23,25^ Anti-myeloperoxidase polyclonal antibody (1:1000 dilution) was coated overnight on a 96-well MediSORP immunoassay plate (ThermoFisher) in buffer consisting of 0.05□JM sodium carbonate/sodium bicarbonate buffer (pH 9.6). After four washes with 0.05% PBS containing Tween-20 (PBS-T), wells were blocked with 2% low-endotoxin BSA (Carl Roth) for 2 hours and subsequently washed another 4 times. Samples were diluted 1:2 and incubated for 90 minutes at room temperature, then washed 4 times before addition of anti-dsDNA-peroxidase antibody from the Cell Death Detection ELISA diluted 1:40 in incubation buffer from the same kit (Roche). Wells were washed another 4 times before development with ready-to-use TMB substrate (Life Technologies, 2023). The reaction was stopped with 1N hydrochloric acid, and the absorbance measured at 450 nM with 630 nM background subtraction using a Gen5 microplate reader (Biotek). A standard concentration curve was prepared from a known amount of MPO standard from the Biolegend myeloperoxidase LEGEND MAX™ ELISA kit pre-incubated with an excess of lambda DNA (Invitrogen) and human native nucleosomes (Merck Millipore), run over a range from 10 ng/ml down to 0.15 ng/ml. All samples measured fell within the limit of quantification based on this standard curve.

##### Statistical Analysis

Data were reported as counts (percentage) or median (interquartile range, IQR). Intergroup differences of baseline characteristics were assessed using the Mann-Whitney test for all continuous variables except for age, which given its normal distribution was assessed using the student t-test. Intergroup differences between categorical variables were assessed using the χ test. Differences in laboratory marker serum concentrations between admission and hospital day 4, pooling results of the entire cohort (unpaired analysis), were assessed using the Mann-Whitney test, and for paired laboratory markers for each individual participant (paired analysis) using the Wilcoxon signed rank test. Cohen’s criteria were used to calculate effect sizes. We performed statistical analyses using SPSS, version 28 (IBM, Armonk, New York). The threshold for statistical significance was set at < 0.05.

##### Data Availability Statement

Data are available on reasonable request from one of the corresponding authors (P.H.).

##### Standard Protocol Approvals, Registrations, and Patients Consents

The study was approved by the Saarland Medical Association ethics committee (#118/17). Written informed consent was obtained from all participants or their legal representatives.

## Results

Among 100 enrolled patients, 83 were found to have an aneurysmal SAH, 5 of which died before follow-up CT scan and were thus ineligible for DCI assessment. In the analytical cohort (n=78), there were 43 (55%) women, and the mean age was 59 years (±13.0) (Table 1). The median World Federation of Neurosurgeons Scale (WFNS) on admission was 2 (IQR, 1-4), and the median modified Fisher scale (mFS) score on admission was 4 (IQR 3-4). Twenty-nine patients (37%) developed DCI. Those who later developed DCI compared to those who did not, had a higher mFS score (median 4 versus 4, p=0.033), and a greater disease severity as illustrated by higher scores on the WFNS (median 3 versus 2, p=0.088), and the Hunt&Hess scale (median 3 versus 2, p=0.061).

**Table 1:** Baseline Characteristics of patients with Aneurysmal Subarachnoid Hemorrhage, Stratified by Presence/Absence of Delayed Cerebral Ischemia.

| **Demographics and medical history** | **All Patients (N=78)** | **DCI (N=29)** | **No DCI (N=49)** | **p-value** |
| --- | --- | --- | --- | --- |
| Mean Age (SD), years | 59.0 (13.0) | 59.7 (12.6) | 58.6 (13.3) | 0.73 |
| Female | 43 (55.1) | 16 (55.2) | 25 (51.0) | 0.995 |
| Hypertension | 41 (52.6) | 16 (55.2) | 25 (51.0) | 0.72 |
| Smoking | 38 (48.7) | 15 (51.7) | 23 (46.9) | 0.68 |
| Coronary artery disease | 9 (11.5) | 4 (13.8) | 5 (10.2) | 0.72 |
| **Clinical Data** |  |  |  |  |
| WFNS score on admission<sup>1</sup> | 2 (1-4.25) | 3 (1-5) | 2 (1-4) | 0.088 |
| WFNS III-V | 29 (37.2) | 15 (51.7) | 19 (38.8) | 0.27 |
| Hunt & Hess score on admission<sup>1</sup> | 3 (2-4) | 3 (2-5) | 2 (2-4) | 0.061 |
| Hunt & Hess IV-V<sup>1</sup> | 25 (37.2) | 12 (41.4) | 13 (26.5) | 0.17 |
| Modified Fisher score on admission<sup>1</sup> | 4 (3-4) | 4 (3-4) | 4 (3-4) | 0.033 |
| Modified Fisher III-V | 65 (83.3) | 27 (93.1) | 38 (77.6) | 0.12 |
| IVH on admission CT | 50 (64.1) | 22 (75.9) | 28 (57.1) | 0.096 |
| ICH on admission CT | 15 (19.2) | 6 (20.7) | 9 (18.4) | 0.80 |
| Hydrocephalus on admission CT | 54 (69.2) | 23 (79.3) | 31 (63.3) | 0.14 |
| Generalized cerebral edema on admission CT | 5 (6.4) | 3 (10.3) | 2 (4.1) | 0.36 |
| Clinical vasospasm | 29 (37.2) | 17 (58.6) | 12 (24.5) | 0.003 |
| Nosocomial infection | 53 (67.9) | 19 (65.5) | 34 (69.4) | 0.72 |
| Admission WBC (x10<sup>3</sup>/μl)<sup>1</sup> | 12.4 (9.6-15.9) | 12.8 (10.7-16.3) | 11.6 (9.4-15.8) | 0.36 |
| **Abbreviations:** DCI, delayed cerebral ischemia; ICH, intracerebral hemorrhage; IVH, intraventricular hemorrhage; mL, milliliters; ng, nanogram; SD, standard deviation; WBC white blood count.<br>P value <0.05 was considered significant<br><sup>1</sup>indicates values presented as median (interquartile range) |  |  |  |  |

### Detection of serum MPO-DNA-complexes

MPO-DNA complexes were detected in all admission and day-4 serum samples. In the pooled unpaired analysis, there was a significant decrease in MPO-DNA levels between admission (median MPO-DNA level 33 ng/ml, IQR, 18-43 ng/ml) and day 4 (median level 22 ng/ml, IQR, 11-31 ng/ml) (Mann-Whitney test, p=0.015). The reduction in MPO-DNA-complex levels coincided with a reduction in white blood counts (WBC) from admission day to day 4 (median 12.4 ×10^3^/μl versus 9.7 ×10^3^/μl, Mann-Whitney test, p<0.001) and an increase in C-reactive protein concentrations (median 2.4, IQR, 1.3-8.3, versus median 174, IQR, 87-254.6, Mann-Whitney-U test p<0.001).

### Primary analysis: MPO-DNA-complexes in patients with and without DCI

In a paired data analysis, there was a significant reduction in MPO-DNA levels between admission and day 4 in patients with DCI (z= -2.095, Wilcoxon signed rank test: p=0.036, r=0.3, medium effect size) but not in those without DCI (Wilcoxon signed rank test: p=0.171) (Figure 1). Like in the unpaired data analysis, there was a significant reduction in WBC in both groups (DCI group p=0.007, no DCI group p<0.001) as well as a significant increase in C-reactive protein (DCI group p<0.001, no DCI group p<0.001) between admission and day 4, respectively.

**Figure 1:**
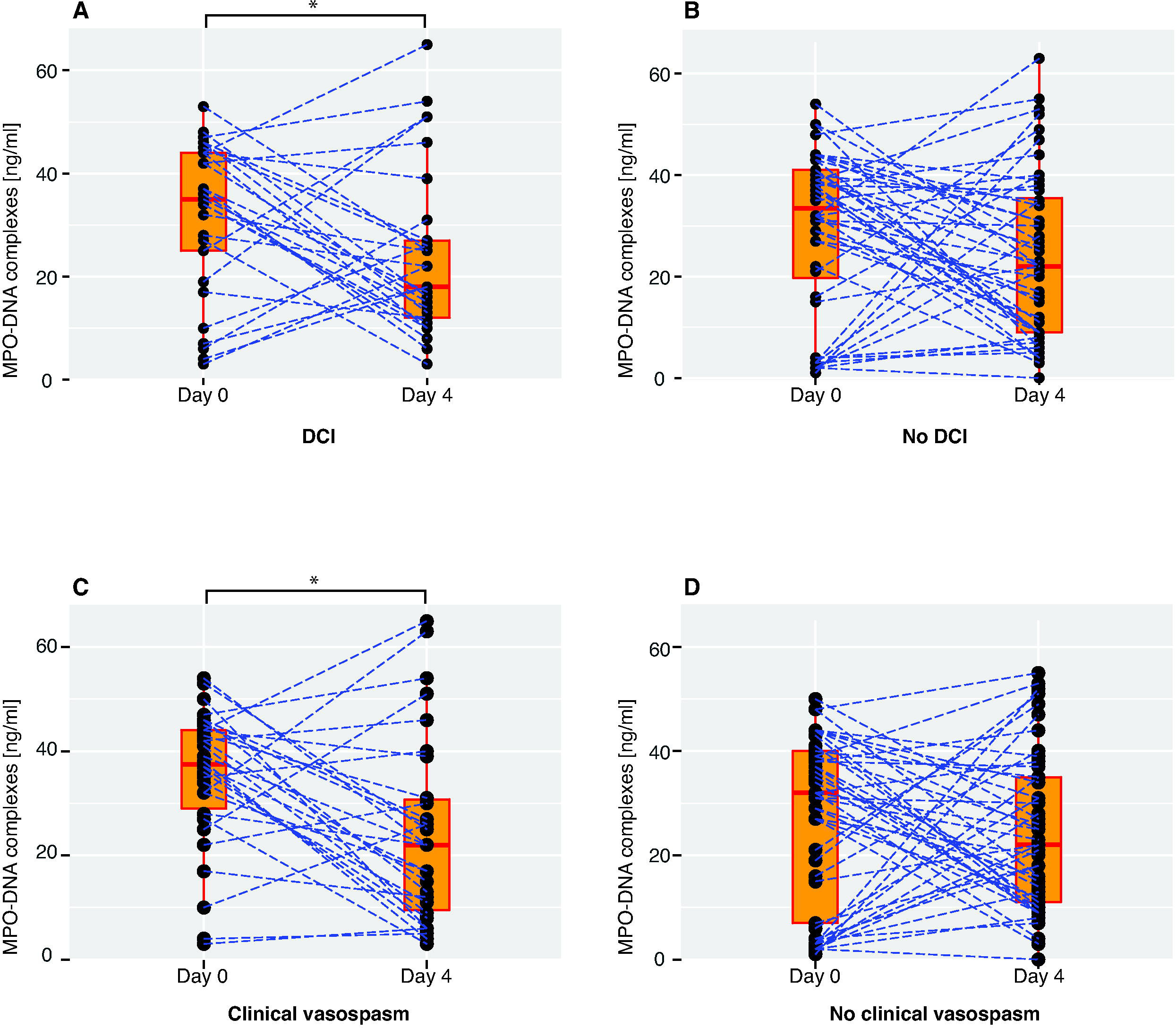
Change in serum MPO-DNA-levels between admission and hospital day 4. Paired data points in the cohort stratified by DCI (present in n=29, absent in n=49, panels A and B), and stratified by clinical vasospasm (present in n=29, absent in n=52, panels C and D). Admission is defined as day 0. Boxes represent median, IQR, and ranges. Dotted blue lines connect paired data points for each participant. Statistical significance (Wilcoxon signed rank test) is indicated by an asterisk. MPO-DNA-complexes: myeloperoxidase-deoxyribonucleic acid-complexes, DCI: delayed cerebral ischemia.

### Secondary analysis: MPO-DNA-complexes in patients with and without clinical vasospasm

When stratifying the cohort by presence or absence of clinical vasospasm, we found a significant reduction in MPO-DNA levels between admission and day 4 in patients with clinical vasospasm (paired analysis, z= -2.757, p=0.006, r=0.4, medium effect size), but not in those without clinical vasospasm (p=0.473) (Figure 1).

## Discussion

NETs, the product of hyperactivated neutrophils, promote immune-mediated thrombus formation and have been associated with ischemic stroke. In this exploratory study in patients with aSAH we detected high levels of MPO-DNA complexes, a NET marker, in all peripheral serum samples. Between admission and hospital day 4, we found a significant reduction in MPO-DNA complex levels in patients who developed DCI, but not in those who did not develop DCI. Our study suggests that NETs might be of relevance in aSAH, possibly contributing to DCI, an ischemic complication strongly associated with poor outcome.

A prior study aimed to detect NET markers in mice and in patients with SAH.^22^ This study used double stranded DNA (ds-DNA) as a surrogate for the presence of NETs. However, ds-DNA has been shown to be an unspecific remnant of cell death^26^. MPO-DNA complexes, in contrast, are established measures of NET release.^17^ Three further studies - in humans, rats, and rabbits respectively - assessed MPO (not MPO-DNA complex) serum levels in SAH.^27-29^ However, secreted MPO levels are indicative of neutrophil activation, including by degranulation, in addition to NET release, which limits the utility of MPO levels in quantifying NETs. Our study adds a novel element to the existing literature, demonstrating the detection of a specific NET biomarker in a prospective cohort of patients with aSAH, as well as its potential implication in the development of DCI.

Among pathophysiologic correlates of DCI, prior studies have primarily investigated vascular dysfunction (leading to vasoconstriction-induced hypoperfusion) and spreading depolarization.^30-32^ In addition, a systemic inflammatory response after SAH, including an increase in the circulating neutrophil-lymphocyte ratio, has been consistently demonstrated across many studies and is associated with DCI and poor outcome.^20,21,33,34^ Disruption of the blood brain barrier with massive influx of cell-free heme into the subarachnoid space may stimulate local macrophages and the heme scavenger system, leading to further recruitment of other immune cells and excessive inflammation.^5,35^ Direct activation of neutrophils through free heme is conceivable as well, with free heme being a driving factor for NET release in sickle cell disease.^36,37^ Whatever the immediate trigger of the inflammatory response to aSAH may be, the concept of NET-induced thrombus formation as a result of it is intriguing, as it provides a framework that links inflammation, thrombus formation, and DCI.^5,38^

We found high levels of MPO-DNA complexes in all patients with aSAH, not only in those with DCI. While lower levels of MPO-DNA complexes have been measured in the general population^39^, the high levels in our study may reflect an overall inflammatory environment after aSAH, in line with high levels of C-reactive protein in all our patients. A speculative explanation for the decline in MPO-DNA complex levels, seen predominantly in patients with DCI, may be NET-driven thrombus formation under consumption of MPO-DNA-complexes. It is worth noting, that using the clinical consensus definition of DCI (which requires diagnosis of an infarct on brain imaging) might be too crude of a parameter to accurately distinguish between patients with and without NET-driven thrombosis, as microthrombosis might not always lead to macroscopic infarcts. This might explain why patients with declining MPO-DNA levels were seen in both the DCI and the no-DCI groups. Demonstrating internal consistency, our results were similar when we stratified the cohort by presence or absence of clinical vasospasm, a more inclusive composite endpoint. This likely reflects previously described associations between DND and DCI on the one hand, and vasospasm and DCI on the other.

### Limitations

Our study has limitations. First, we measured MPO-DNA complex levels in serum. In contrast to plasma, serum preparations may result in the release of NETs during the coagulation phase and may thus artificially increase NET levels. The advantage of serum, however, is that NET markers can be paired with neutrophil counts to infer the extent of NET release during serum preparation due to neutrophil preactivation in the collected blood samples (which is not possible in plasma samples because neutrophils are removed). In this study, matched complete blood counts were not available in our patient cohort, hence, correlation of MPO-DNA complexes and absolute neutrophil counts could not be performed. Second, due to the exploratory character of this study, we refrained from analyzing NET markers other than MPO-DNA complexes. Third, we did not analyze samples beyond hospital day 4. Since DCI typically occurs between post-SAH day 4 to 14, it is possible that in those patients with DCI, further reductions in MPO-DNA complex levels might have been appreciated at later time points. Future studies should ideally use paired NET measurements of plasma and serum samples with matching complete blood counts and include measurements of further NET biomarkers with additional mechanistic insight, e.g., citrullinated histones and peptidylarginine deiminase 4.

## Conclusions

MPO-DNA complexes, a component of NETs, are detectable in the peripheral serum of patients with aSAH. MPO-DNA complex levels undergo a marked reduction in those with DCI over the first days of hospitalization, suggesting a potential role of NETs in the pathophysiology of DCI.

## Data Availability

Data are available on reasonable request from one of the corresponding authors (P.H.).

## Acknowledgments

None.

## References

1. Muehlschlegel S. Subarachnoid Hemorrhage. Continuum (Minneap Minn). 2018;24:1623–1657. doi: 10.1212/CON.0000000000000679

2. Witsch J, Frey HP, Patel S, Park S, Lahiri S, Schmidt JM, Agarwal S, Falo MC, Velazquez A, Jaja B, et al. Prognostication of long-term outcomes after subarachnoid hemorrhage: The FRESH score. Ann Neurol. 2016;80:46–58. doi: 10.1002/ana.24675

3. Vergouwen MD, Vermeulen M, van Gijn J, Rinkel GJ, Wijdicks EF, Muizelaar JP, Mendelow AD, Juvela S, Yonas H, Terbrugge KG, et al. Definition of delayed cerebral ischemia after aneurysmal subarachnoid hemorrhage as an outcome event in clinical trials and observational studies: proposal of a multidisciplinary research group. Stroke. 2010;41:2391–2395. doi: 10.1161/strokeaha.110.589275

4. Al-Mufti F, Roh D, Lahiri S, Meyers E, Witsch J, Frey HP, Dangayach N, Falo C, Mayer SA, Agarwal S, et al. Ultra-early angiographic vasospasm associated with delayed cerebral ischemia and infarction following aneurysmal subarachnoid hemorrhage. J Neurosurg. 2017;126:1545–1551. doi: 10.3171/2016.2.JNS151939

5. Dodd WS, Laurent D, Dumont AS, Hasan DM, Jabbour PM, Starke RM, Hosaka K, Polifka AJ, Hoh BL, Chalouhi N. Pathophysiology of Delayed Cerebral Ischemia After Subarachnoid Hemorrhage: A Review. J Am Heart Assoc. 2021;10:e021845. doi: 10.1161/JAHA.121.021845

6. Sarrafzadeh AS, Vajkoczy P, Bijlenga P, Schaller K. Monitoring in Neurointensive Care - The Challenge to Detect Delayed Cerebral Ischemia in High-Grade Aneurysmal SAH. Frontiers in neurology. 2014;5:134. doi: 10.3389/fneur.2014.00134

7. Megjhani M, Terilli K, Weiss M, Savarraj J, Chen LH, Alkhachroum A, Roh DJ, Agarwal S, Connolly ES, Jr., Velazquez A, et al. Dynamic Detection of Delayed Cerebral Ischemia: A Study in 3 Centers. Stroke. 2021;52:1370–1379. doi: 10.1161/STROKEAHA.120.032546

8. Chen HY, Elmer J, Zafar SF, Ghanta M, Junior VM, Rosenthal ES, Gilmore EJ, Hirsch LJ, Zaveri HP, Sheth KN, et al. Combining Transcranial Doppler and EEG Data to Predict Delayed Cerebral Ischemia After Subarachnoid Hemorrhage. Neurology. 2021. doi: 10.1212/wnl.0000000000013126

9. Lissak IA, Locascio JJ, Zafar SF, Schleicher RL, Patel AB, Leslie-Mazwi T, Stapleton CJ, Koch MJ, Kim JA, Anderson K, et al. Electroencephalography, Hospital Complications, and Longitudinal Outcomes After Subarachnoid Hemorrhage. Neurocrit Care. 2021;35:397–408. doi: 10.1007/s12028-020-01177-x

10. Jabbarli R, Pierscianek D, Darkwah Oppong M, Sato T, Dammann P, Wrede KH, Kaier K, Kohrmann M, Forsting M, Kleinschnitz C, et al. Laboratory biomarkers of delayed cerebral ischemia after subarachnoid hemorrhage: a systematic review. Neurosurg Rev. 2018. doi: 10.1007/s10143-018-1037-y

11. Martinod K, Wagner DD. Thrombosis: tangled up in NETs. Blood. 2014;123:2768–2776. doi: 10.1182/blood-2013-10-463646

12. Brinkmann V, Reichard U, Goosmann C, Fauler B, Uhlemann Y, Weiss DS, Weinrauch Y, Zychlinsky A. Neutrophil extracellular traps kill bacteria. Science (New York, NY). 2004;303:1532–1535. doi: 10.1126/science.1092385

13. Laridan E, Denorme F, Desender L, Francois O, Andersson T, Deckmyn H, Vanhoorelbeke K, De Meyer SF. Neutrophil extracellular traps in ischemic stroke thrombi. Ann Neurol. 2017;82:223–232. doi: 10.1002/ana.24993

14. Martinod K, Witsch T, Farley K, Gallant M, Remold-O’Donnell E, Wagner DD. Neutrophil elastase-deficient mice form neutrophil extracellular traps in an experimental model of deep vein thrombosis. J Thromb Haemost. 2016;14:551–558. doi: 10.1111/jth.13239

15. Blasco A, Coronado MJ, Hernandez-Terciado F, Martin P, Royuela A, Ramil E, Garcia D, Goicolea J, Del Trigo M, Ortega J, et al. Assessment of Neutrophil Extracellular Traps in Coronary Thrombus of a Case Series of Patients With COVID-19 and Myocardial Infarction. JAMA Cardiol. 2020. doi: 10.1001/jamacardio.2020.7308

16. Papayannopoulos V. Neutrophil extracellular traps in immunity and disease. Nat Rev Immunol. 2018;18:134–147. doi: 10.1038/nri.2017.105

17. Sorvillo N, Cherpokova D, Martinod K, Wagner DD. Extracellular DNA NET-Works With Dire Consequences for Health. Circ Res. 2019;125:470–488. doi: 10.1161/circresaha.119.314581

18. Pena-Martinez C, Duran-Laforet V, Garcia-Culebras A, Ostos F, Hernandez-Jimenez M, Bravo-Ferrer I, Perez-Ruiz A, Ballenilla F, Diaz-Guzman J, Pradillo JM, et al. Pharmacological Modulation of Neutrophil Extracellular Traps Reverses Thrombotic Stroke tPA (Tissue-Type Plasminogen Activator) Resistance. Stroke. 2019;50:3228–3237. doi: 10.1161/STROKEAHA.119.026848

19. Ducroux C, Di Meglio L, Loyau S, Delbosc S, Boisseau W, Deschildre C, Ben Maacha M, Blanc R, Redjem H, Ciccio G, et al. Thrombus Neutrophil Extracellular Traps Content Impair tPA-Induced Thrombolysis in Acute Ischemic Stroke. Stroke. 2018;49:754–757. doi: 10.1161/STROKEAHA.117.019896

20. Claassen J, Albers D, Schmidt JM, De Marchis GM, Pugin D, Falo CM, Mayer SA, Cremers S, Agarwal S, Elkind MS, et al. Nonconvulsive seizures in subarachnoid hemorrhage link inflammation and outcome. Ann Neurol. 2014;75:771–781. doi: 10.1002/ana.24166

21. Al-Mufti F, Amuluru K, Damodara N, Dodson V, Roh D, Agarwal S, Meyers PM, Connolly ES, Jr., Schmidt MJ, Claassen J, et al. Admission neutrophil-lymphocyte ratio predicts delayed cerebral ischemia following aneurysmal subarachnoid hemorrhage. J Neurointerv Surg. 2019;11:1135–1140. doi: 10.1136/neurintsurg-2019-014759

22. Früh A, Tielking K, Schoknecht F, Liu S, Schneider UC, Fischer S, Vajkoczy P, Xu R. RNase A Inhibits Formation of Neutrophil Extracellular Traps in Subarachnoid Hemorrhage. Frontiers in Physiology. 2021;12. doi: 10.3389/fphys.2021.724611

23. Hemmer S, Senger S, Griessenauer CJ, Simgen A, Oertel J, Geisel J, Hendrix P. Admission serum high mobility group box 1 (HMGB1) protein predicts delayed cerebral ischemia following aneurysmal subarachnoid hemorrhage. Neurosurg Rev. 2021. doi: 10.1007/s10143-021-01607-0

24. de Oliveira Manoel AL, van der Jagt M, Amin-Hanjani S, Bambakidis NC, Brophy GM, Bulsara K, Claassen J, Connolly ES, Hoffer SA, Hoh BL, et al. Common Data Elements for Unruptured Intracranial Aneurysms and Aneurysmal Subarachnoid Hemorrhage: Recommendations from the Working Group on Hospital Course and Acute Therapies-Proposal of a Multidisciplinary Research Group. Neurocrit Care. 2019;30:36–45. doi: 10.1007/s12028-019-00726-3

25. Vanderbeke L, Van Mol P, Van Herck Y, De Smet F, Humblet-Baron S, Martinod K, Antoranz A, Arijs I, Boeckx B, Bosisio FM, et al. Monocyte-driven atypical cytokine storm and aberrant neutrophil activation as key mediators of COVID-19 disease severity. Nat Commun. 2021;12:4117. doi: 10.1038/s41467-021-24360-w

26. Langseth MS, Helseth R, Ritschel V, Hansen CH, Andersen GO, Eritsland J, Halvorsen S, Fagerland MW, Solheim S, Arnesen H, et al. Double-Stranded DNA and NETs Components in Relation to Clinical Outcome After ST-Elevation Myocardial Infarction. Sci Rep. 2020;10:5007. doi: 10.1038/s41598-020-61971-7

27. Lim M, Bower RS, Wang Y, Sims L, Bower MR, Camara-Quintana J, Li G, Cheshier S, Harsh GRt, Steinberg GK, et al. The predictive value of serum myeloperoxidase for vasospasm in patients with aneurysmal subarachnoid hemorrhage. Neurosurg Rev. 2012;35:413–419; discussion 419. doi: 10.1007/s10143-012-0375-4

28. Ersahin M, Toklu HZ, Erzik C, Cetinel S, Akakin D, Velioglu-Ogunc A, Tetik S, Ozdemir ZN, Sener G, Yegen BC. The anti-inflammatory and neuroprotective effects of ghrelin in subarachnoid hemorrhage-induced oxidative brain damage in rats. J Neurotrauma. 2010;27:1143–1155. doi: 10.1089/neu.2009.1210

29. Zhou ML, Shi JX, Hang CH, Cheng HL, Qi XP, Mao L, Chen KF, Yin HX. Potential contribution of nuclear factor-kappaB to cerebral vasospasm after experimental subarachnoid hemorrhage in rabbits. J Cereb Blood Flow Metab. 2007;27:1583–1592. doi: 10.1038/sj.jcbfm.9600456

30. Saber H, Desai A, Palla M, Mohamed W, Seraji-Bozorgzad N, Ibrahim M. Efficacy of Cilostazol in Prevention of Delayed Cerebral Ischemia after Aneurysmal Subarachnoid Hemorrhage: A Meta-Analysis. J Stroke Cerebrovasc Dis. 2018;27:2979–2985. doi: 10.1016/j.jstrokecerebrovasdis.2018.06.027

31. Macdonald RL, Higashida RT, Keller E, Mayer SA, Molyneux A, Raabe A, Vajkoczy P, Wanke I, Bach D, Frey A, et al. Clazosentan, an endothelin receptor antagonist, in patients with aneurysmal subarachnoid haemorrhage undergoing surgical clipping: a randomised, double-blind, placebo-controlled phase 3 trial (CONSCIOUS-2). Lancet Neurol. 2011;10:618–625. doi: 10.1016/S1474-4422(11)70108-9

32. Dreier JP, Major S, Manning A, Woitzik J, Drenckhahn C, Steinbrink J, Tolias C, Oliveira-Ferreira AI, Fabricius M, Hartings JA, et al. Cortical spreading ischaemia is a novel process involved in ischaemic damage in patients with aneurysmal subarachnoid haemorrhage. Brain. 2009;132:1866–1881. doi: 10.1093/brain/awp102

33. Tam AK, Ilodigwe D, Mocco J, Mayer S, Kassell N, Ruefenacht D, Schmiedek P, Weidauer S, Pasqualin A, Macdonald RL. Impact of systemic inflammatory response syndrome on vasospasm, cerebral infarction, and outcome after subarachnoid hemorrhage: exploratory analysis of CONSCIOUS-1 database. Neurocrit Care. 2010;13:182–189. doi: 10.1007/s12028-010-9402-x

34. Badjatia N, Monahan A, Carpenter A, Zimmerman J, Schmidt JM, Claassen J, Connolly ES, Mayer SA, Karmally W, Seres D. Inflammation, negative nitrogen balance, and outcome after aneurysmal subarachnoid hemorrhage. Neurology. 2015;84:680–687. doi: 10.1212/WNL.0000000000001259

35. Hugelshofer M, Buzzi RM, Schaer CA, Richter H, Akeret K, Anagnostakou V, Mahmoudi L, Vaccani R, Vallelian F, Deuel JW, et al. Haptoglobin administration into the subarachnoid space prevents hemoglobin-induced cerebral vasospasm. J Clin Invest. 2019;129:5219–5235. doi: 10.1172/JCI130630

36. Graca-Souza AV, Arruda MA, de Freitas MS, Barja-Fidalgo C, Oliveira PL. Neutrophil activation by heme: implications for inflammatory processes. Blood. 2002;99:4160–4165. doi: 10.1182/blood.v99.11.4160

37. Chen G, Zhang D, Fuchs TA, Manwani D, Wagner DD, Frenette PS. Heme-induced neutrophil extracellular traps contribute to the pathogenesis of sickle cell disease. Blood. 2014;123:3818–3827. doi: 10.1182/blood-2013-10-529982

38. Goursaud S, Martinez de Lizarrondo S, Grolleau F, Chagnot A, Agin V, Maubert E, Gauberti M, Vivien D, Ali C, Gakuba C. Delayed Cerebral Ischemia After Subarachnoid Hemorrhage: Is There a Relevant Experimental Model? A Systematic Review of Preclinical Literature. Front Cardiovasc Med. 2021;8:752769. doi: 10.3389/fcvm.2021.752769

39. Donkel SJ, Wolters FJ, Ikram MA, de Maat MPM. Circulating Myeloperoxidase (MPO)-DNA complexes as marker for Neutrophil Extracellular Traps (NETs) levels and the association with cardiovascular risk factors in the general population. PLoS One. 2021;16:e0253698. doi: 10.1371/journal.pone.0253698

